# Mathematical modelling of dynamics and containment of COVID-19 in Ukraine

**DOI:** 10.1101/2020.07.24.20161497

**Authors:** Yuliya N. Kyrychko, Konstantin B. Blyuss, Igor Brovchenko

## Abstract

COVID-19 disease caused by the novel SARS-Cov-2 coronavirus has already brought unprecedented challenges for public health and resulted in huge numbers of cases and deaths worldwide. In the absence of effective vaccine, different countries have employed various other types of non-pharmaceutical interventions to contain the spread of this disease, including quarantines and lockdowns, tracking, tracing and isolation of infected individuals, and social distancing measures. Effectiveness of these and other measures of disease containment and prevention to a large degree depends on good understanding of disease dynamics, and robust mathematical models play an important role for forecasting its future dynamics. In this paper we focus on Ukraine, one of Europe’s largest countries, and develop a mathematical model of COVID-19 dynamics, using latest data on parameters characterising clinical features of disease. For improved accuracy, our model includes age-stratified disease parameters, as well as age- and location-specific contact matrices to represent contacts. We show that the model is able to provide an accurate short-term forecast for the numbers and age distribution of cases and deaths. We also simulated different lockdown scenarios, and the results suggest that reducing work contacts is more efficient at reducing the disease burden than reducing school contacts, or implementing shielding for people over 60.

## Introduction

The first registered cases of novel COVID-19 disease appeared in mid-December 2019 in Wuhan, China^1^, and due to human-to-human transmission, the number of infected people grew rapidly forcing Wuhan to go into a strict lockdown^2^. Since then, the disease was declared a global pandemic by the WHO on the 11th March 2020, and has quickly spread all around the world, infecting populations in all countries and taking numerous lives in its course. In order to stop the spread of this deadly infection and in the absence of effective disease-specific antiviral treatment or vaccine, many countries have followed China and introduced lockdowns, closing borders and stopping all usual day-to-day activities, such as closures of schools, work places and similar. The introduction of social distancing measures analysed using mathematical models has shown their effectiveness in reducing the spread of COVID-19 infection^3–9^.

The COVID-19 infection is mainly transmitted via respiratory route, and the main symptoms are dry cough, high fever, and loss of smell and/or taste. Some carriers are completely asymptomatic, and estimates suggest the number of asymptomatic individuals can be as high as 50-75%^10,11^. The disease severity varies depending on age, and most deaths occur in people over the age of 65, or in those with pre-existing conditions. The challenging factors in managing the spread of COVID-19 include its high infectivity, and the most severe cases requiring intensive care treatment often being accompanied by the need in ventilator stimulation^12,13^. The incubation period for COVID-19 is about 5.5 days, but can be up to 14 days, and this varies between different individuals^14^. Recent analysis suggests that since the viral loads of symptomatic and asymptomatic individuals are similar, the asymptomatic carriers have the same ability to infect^15^.

The first time Ukraine registered more than 100 people infected with the novel COVID-19 disease was on the 25th March 2020, with the majority of infected cases coming from abroad, and the early COVID-19 progression was similar to that of Sweden and Poland, albeit with some delay. To halt the spread of the COVID-19, Ukraine went into lockdown on the 17th March 2020, and had 14 confirmed cases and 2 deaths the following day, with more strict restrictions being introduced on the 6th April 2020, including the closure of schools, universities. shopping malls, fitness facilities. Public transport was reduced to an absolute minimum across the country to minimise inter-regional transmissions, and face masks became obligatory in all public places. The early introduction of lockdown slowed the disease progression, with estimated doubling time of cases going from 8 days to 11 days in the week 28th April - 5th May 2020, and during this time, the most infected regions, such as Kyiv and Chernivetska region showed the decrease in doubling of infections to 10 days. The following week, the doubling of COVID-19 cases across Ukraine increased from 11 to 13 days, and was showing a very rapid slowing down, provided lockdown measures are observed. The first easing of lockdown started on the 12th May 2020 with lifting of further restrictions on the 22nd May 2020. As of 10th July 2020, there are 52, 043 confirmed cases of COVID-19, and 1, 345 deaths in Ukraine, with the number of confirmed cases steadily and significantly increasing every day since the middle of June 2020, and then slowing down in the first two weeks of July 2020.

In this paper we consider the dynamics of COVID-19 epidemic in Ukraine, and model it using a modified age-structured compartmental SEIR (susceptible, exposed, infected and recovered) framework. The modifications account for the presence of asymptomatic carriers, different types of clinical progression of the disease once someone becomes infected, the need for hospitalisation, as well as the severity of the hospital interventions. The model assumes that individuals who have recovered from COVID-19, subsequently have immunity against the virus for the remaining duration of the epidemic. Official data from the Ministry of Health of Ukraine regarding confirmed daily infected cases and deaths, as well as distributions of incubation and recovery times, are used to parameterise the model. We also provide a short-term forecast of further dynamics of COVID-19 in Ukraine and explore the impact of different types of lockdown on the number of cases and deaths.

## Results

### Mathematical model

We have developed a compartmental SEIR-type model that accounts for specific distributions of different characteristic times, as well as the population structure in the country. This model extends an earlier work of Blyuss and Kyrychko^16^ on COVID-19 dynamics in the UK and that of Brovchenko^17^ on mean-field COVID-19 dynamics in Ukraine. The total population is divided into the following groups: susceptible individuals to be denoted as *S*(*t*), exposed individuals *E*(*t*), infected individuals *I*(*t*). We assume that only infected individuals are capable of transmitting the infection to susceptibles, and we also assume that after the end of infectious period, a proportion *p*_*A*_ of infected individuals (asymptomatic carriers) will simply recover without ever exhibiting any symptoms of the disease, thus moving to a compartment of asymptomatic recovered individuals *R*_*A*_(*t*), and similarly a proportion *p*_*M*_ of individuals with mild symptoms will also proceed to a recovered class *R*_*M*_(*t*). Following Davies et al.^18^ (see also^19–21^), we further assume that after the same period of time, a proportion *p*_*H*_ of individuals will require hospitalisation. Among these individuals, a proportion *p*_*S*_ move to a new compartment *I*_*H*_(*t*) containing individuals with severe disease, who will recover from infection in hospital, and upon discharge will move to a recovered compartment *R*_*H*_(*t*). The remaining proportion *p*_*F*_ will spend a period of time in hospital, and will eventually die, thus moving to a compartment *D*(*t*). The diagram of transitions between different model compartments is shown in Fig. 1, and the model itself is presented in the *Methods* section.

**Figure 1.**
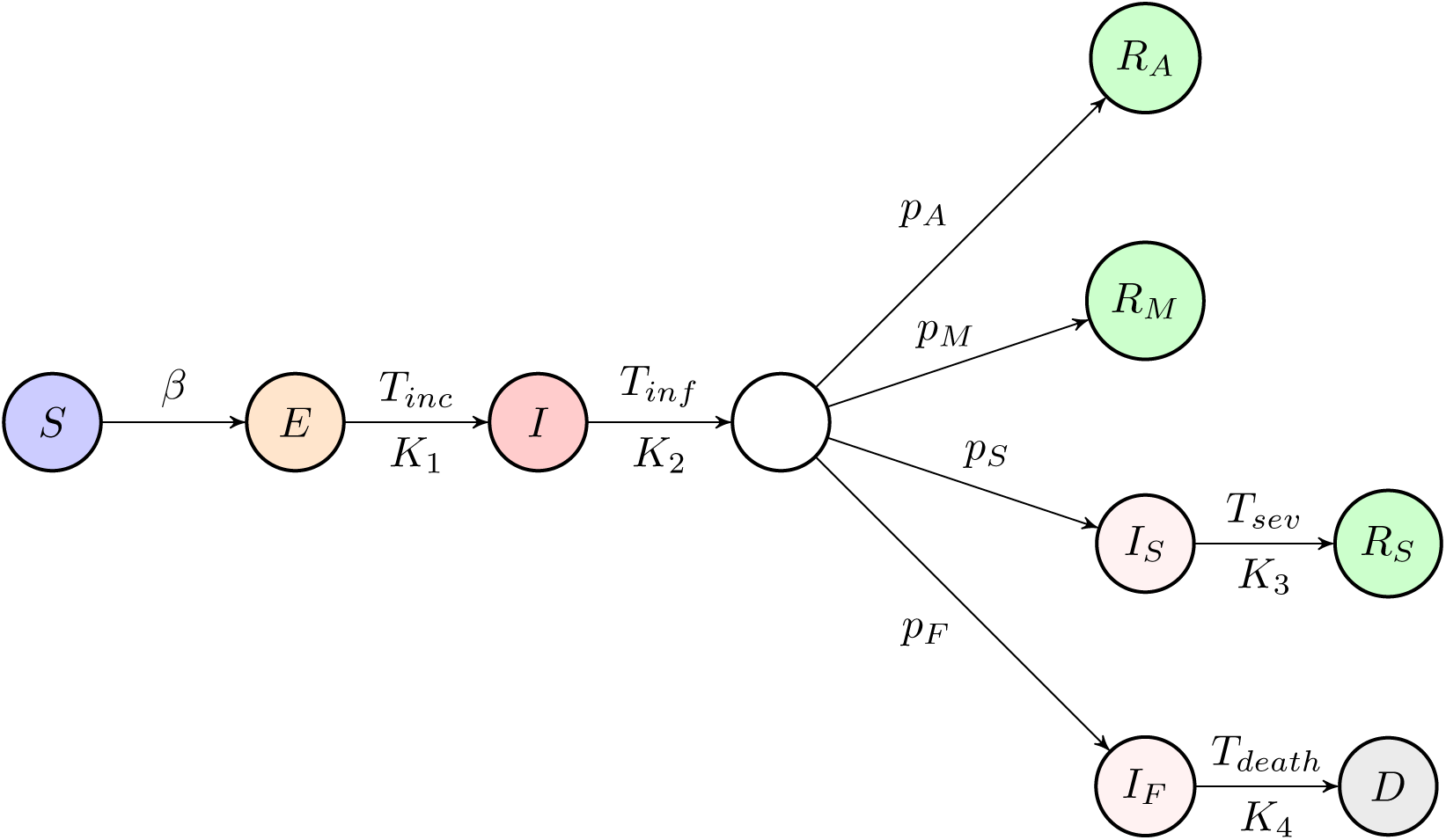
Schematic diagram of the disease transmission model.

Analysis of data collected by the Public Health Center of the Ministry of Health of Ukraine on distributions of incubation period *T*_*inc*_, the period of recovery and discharge from hospital for severe cases *T*_*sev*_, and the time to death in hospital *T*_*death*_, shows that they appear to be well-approximated by a gamma distribution, as illustrated in Fig. 2. Similar distributions for incubation and infectious periods have been obtained in the studies of epidemiological data for COVID-19 cases in other countries^22,23^. In the model, we represent these distributions in the form of multiple stages with the same mean period^24,25^. In the absence of clinical data on the distribution of infectious period *T*_*in f*_, it was taken to obey a gamma distribution with the mean duration of 5 days and four stages, as used in a recent work of Davies et al.^18^.

**Figure 2.**
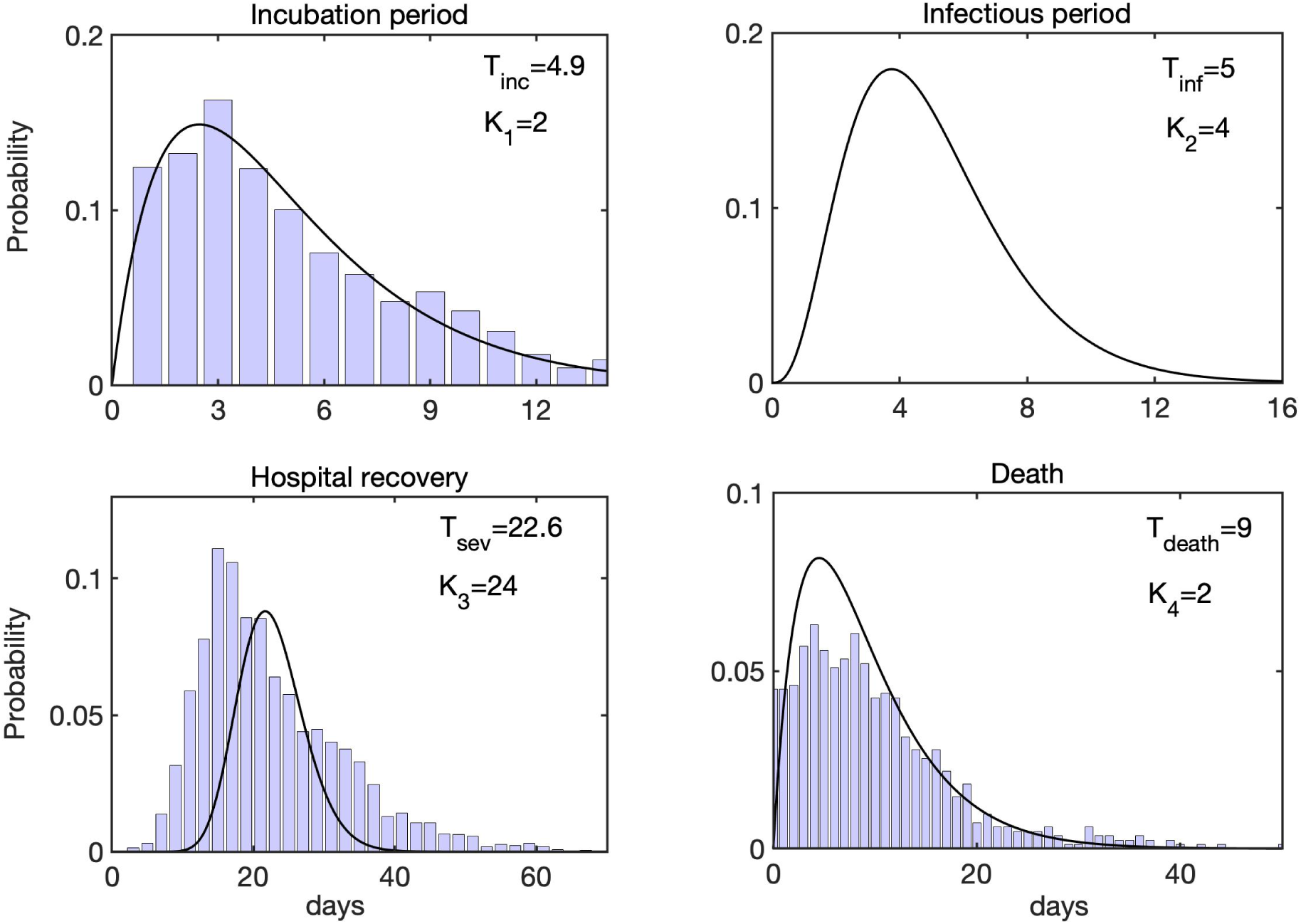
Distributions of incubation time, infectious period, hospital recovery period, and time to death. Histograms show actual data from^47^, black curves are fitted gamma distributions with the corresponding parameters.

Since population structure is known to have a major effect on disease transmission, as manifested by significant differences in age-specific hospitalisation and mortality rates, we have divided each compartment into sixteen 5-year age groups spanning from 0 until 75+. To model interactions between individuals in different age groups, we have used a contact matrix

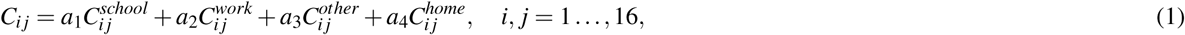

which includes contributions from individual contact matrices representing the numbers of daily contacts between individuals in different settings, namely, school, work, home, and other. Initially, we take *a*_1_ = *a*_2_ = *a*_3_ = *a*_4_ = 1, and by varying these rates one can model the effects of different intervention strategies^18,26,27^. Age-specific data on the percentage of asymptomatic cases, as well as hospitalisation and mortality are given in Table 1 in the *Methods* section.

**Table 1.** Age-specific parameter values^50^.

| Age-group (years) | % asymptomatic cases, $p_A$ | % hospitalised cases, $p_H$ | case fatality ratio, $p_F$ |
| --- | --- | --- | --- |
| 0 to 4 | 45.8% | 19.3% | 0.002% |
| 5 to 9 | 46.9% | 12.4% | 0.002% |
| 10 to 14 | 46.3% | 10.5% | 0.10% |
| 15 to 19 | 43.1% | 13.3% | 0.34% |
| 20 to 24 | 42.3% | 14.6% | 0.11% |
| 25 to 29 | 39.3% | 14.4% | 0.11% |
| 30 to 34 | 40% | 15.2% | 0.36% |
| 35 to 39 | 36.4% | 17% | 0.43% |
| 40 to 44 | 35.6% | 21.7% | 0.92% |
| 45 to 49 | 35.5% | 23.5% | 1.27% |
| 50 to 54 | 33.2% | 28.3% | 1.76% |
| 55 to 59 | 30.2% | 33.3% | 2.77% |
| 60 to 64 | 28.7% | 36.7% | 3.70% |
| 65 to 69 | 28.5% | 41.6% | 5.95% |
| 70 to 74 | 28.2% | 45.5% | 8.44% |
| 75 to 79 | 26.4% | 50.8% | 11.25% |
| 80+ | 27% | 52.9% | 18.2% |

### Dynamics of epidemic spread

To model the dynamics of the spread of SARS-Cov-2 in Ukraine, we took into account various containment measures that were introduced at different times, and incorporated these in the model by changing the values of coefficients *a*_1_, …, *a*_4_ for different contributions to a mixing matrix (1), as has been done in a number of recent models of COVID-19 in other countries^18,26,27^. This also fits with early observations on the effects of different intervention measures on reducing the numbers of cases in Hubei province where the first cases of SARS-Cov-2 virus were registered^28^. The start of simulation, which corresponds to 25 March 2020, coincides with the date when Ukraine went into a state of lockdown. By this point, schools and universities closed, which we modelled by setting *a*_1_ = 0, and similarly nurseries and playgrounds also closed at that date, which we represented by setting 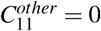. The other coefficients were set as *a*_1_ = *a*_2_ = 0.4 (significantly reduced interactions in ‘work’ and ‘other’ compartments), whereas *a*_4_ has remained equal to one throughout. From the 6th April 2020, new governmental guidance introduced shielding for self-isolation for over-60s, hence we reduced their entries in ‘work’ and ‘other’ matrices by another 50%. All these restrictions remained in force until 25 May 2020, when nurseries opened, but with social distancing requirements, and from the 1st June 2020, gyms, schools and universities were also allowed to open for very limited service, which we represent by setting *a*_1_ = 1 and changing 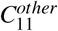 to 30% of its baseline value. Finally, on the 5th June 2020, a restriction on self-isolation of over-60s was lifted, together with limited opening of cafes/restaurants etc., for a subsequent simulation the values of all coefficients were *a*_1_ = 0.1, *a*_2_ = *a*_3_ = 0.6, *a*_4_ = 1.

Figure 3 shows the results of numerical simulations of the model (2) with parameter values from Table 1. It demonstrates a good agreement between short-term predictions, obtained using the model, with actual data, even despite all the challenges associated with accurate and timely collection and reporting of cases and deaths. In terms of dynamics, one observes an extended plateau-like region, starting from the middle of April 2020 and lasting until the end of May 2020. Whereas the total numbers of cases and deaths are monotonically increasing, the numbers of daily new cases and deaths have remained largely unchanged during this period. Epidemiologically, this can be explained by the fact that unlike many other countries, Ukraine rather went into a lockdown already in the middle of March 2020, when the number of confirmed cases was still very small, and the quarantine has largely remained in place until May. However, two months later it became harder to justify maintenance of strict lockdown rules, and they started to be relaxed. This has included allowing people over 60 years old to leave their homes (before that they were expected to be shielding and not leave home except for emergency), resuming operation of public transport, as well as resumption of work in cafes and restaurants, gyms etc. As the lockdown restriction were lifted, this led to a significant increase in mixing between people, with many not following social distancing guidelines, and, similarly to what has been observed in many other countries, the numbers of cases and deaths have started to grow significantly. One particular challenge is that in many Ukrainian towns and cities, transportation is delivered mostly by shuttle taxis, and due to a limited number of these taxis, they often take large numbers of people, which effectively results in people being in very close quarters for an extended period of time without good ventilation, which facilitates SARS-Cov-2 airborne transmission. Latest figures from the beginning of July indicate that existing social distancing measures appear to be working, with the growth in new cases reducing over this period.

**Figure 3.**
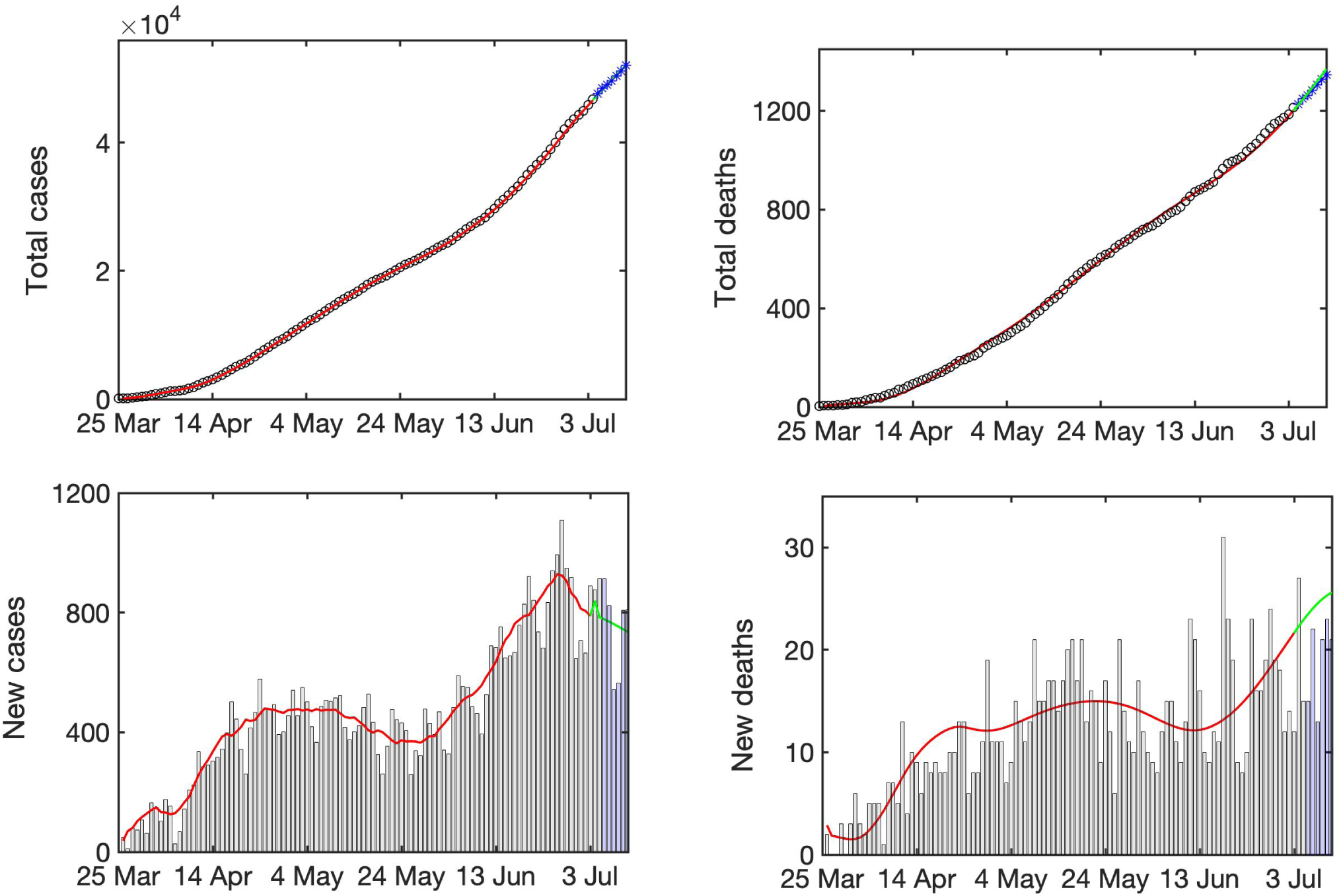
Total numbers of cases and deaths, and the daily numbers of cases and deaths. Black circles (black histograms) show actual data, red line shows simulation of the model (2), green line shows a projection using simulation of model (2), blue stars (blue histograms) show latest actual data. All data are taken from^48^.

In Fig. 4 we illustrate age distributions for cases and deaths as observed on the 10 July 2020. Similarly to what has been observed in other countries^18,23^, the numbers of deaths are small among younger people and generally increase with age. Quite worryingly, the proportion of deaths is quite large already for people of 50-60 years of age (around 20%), and the highest in the age bracket of 60-70 years (30%), with smaller proportions in older people (24% in 70-80, and 17% in 80+). One possible explanation for this is the fact that the average life expectancy in Ukraine is currently 66.6 years for males and 76.7 for females^29^, which suggests that people in 60-70 age group are very likely to die from a variety of causes, with COVID-19 possibly just exacerbating this. The fact that the proportion of deaths among the 80+ group is smaller may be attributed to the fact that while this age group has the highest age-specific case fatality ratio, they are also the smallest in terms of actual numbers of people. Another really worrying trend is the fact that deaths among the 40-50 age group amount to around 7% of total deaths, whereas in many other countries the proportion of deaths in this age group is much smaller. This could potentially be attributed to co-morbidities among people in this age group that result in increasing the death rate. In terms of the numbers of confirmed cases, working-age categories of people appear to bear the highest disease burden, though around 10% of cases have been registered in children and young adults under 20. While the numbers of deaths among children are very low, they are still non-negligible, and some recent outbreaks in nurseries and schools highlight the importance of maintaining social distancing in these settings, where naturally there is a high degree mixing between children. Importantly, for both cases and deaths, the age distribution essentially stabilised by mid-April, and subsequent growth of cases and deaths does not appear to change the age distribution, so cases and deaths in each specific age groups are just proportionally growing. One should note that despite qualitative similarities, there are some discrepancies between simulated and observed age distributions of cases and deaths shown in Fig. 4. This can be attributed to a variety of factors: the mixing matrices used for simulations were inferred from a POLYMOD matrix using household survey data, but they may not completely accurately represent actual mixing patterns in Ukrainian population; there are notable differences in population structure and mixing in different regions of Ukraine; there are certain issues with collecting and reporting of cases and deaths. Despite all these challenges for accurate predictions, our model provides important quantitative insights into COVID-19 dynamics, particularly from the perspective of impact of different age groups and population structure on disease spread.

**Figure 4.**
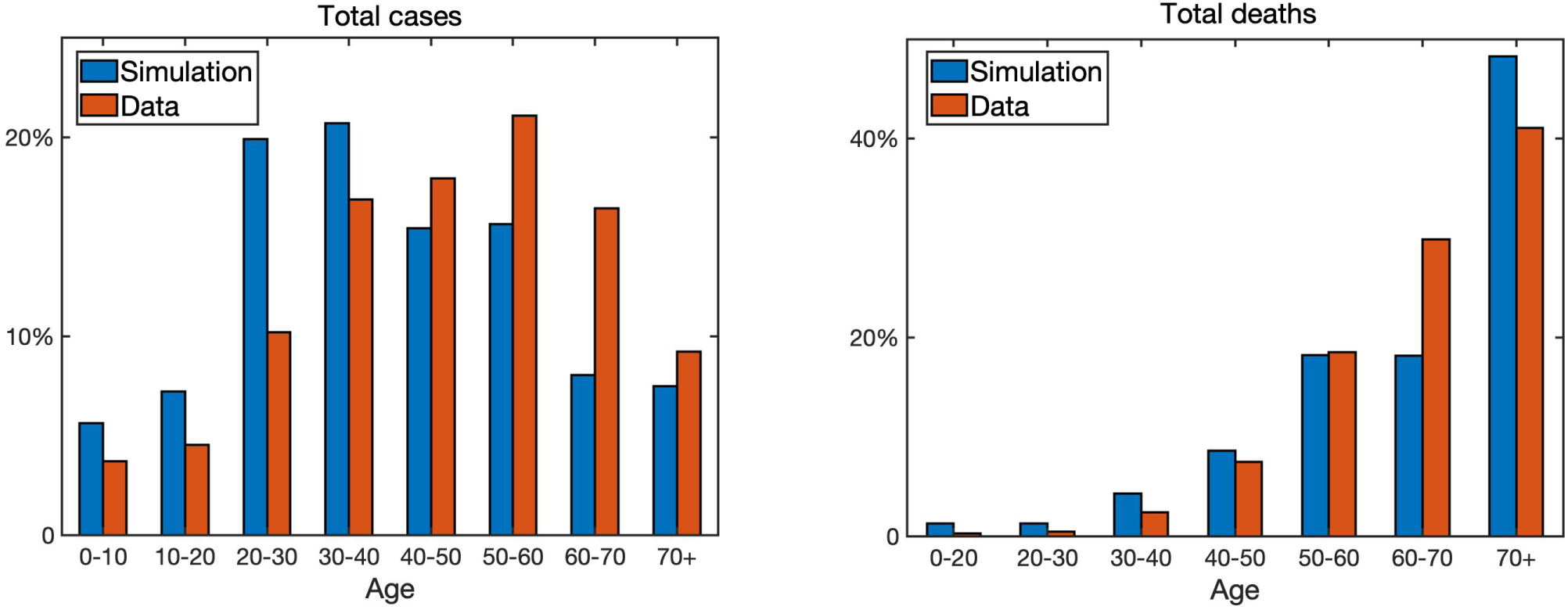
Age distribution of COVID-19 cases and deaths on the 10 July 2020 from the data^48^ and from the simulation of model (2).

### Longer-term forecast and containment

Figure 5 compares longer-term forecasts of epidemic dynamics for the baseline value of disease transmission rate fixed at the value it reached on the 10 July 2020, with scenarios when the transmission rate is increased uniformly for all age groups by 10% or 20% over a period of two weeks. The downward trend in the number of new cases observed in the first ten days of July results in the continuing reduction in the number of new cases and deaths (with deaths slightly lagging), and slowing the growth rates of total cases and deaths. An increase of disease transmission for all age groups by 10% results in raising the effective reproduction number, and with the planned resumption of studies in schools from the 1st September 2020 (though with social distancing measures in place, so we set *a*_1_ = 0.6), this is sufficient to result in a reversal of dynamics and the growth in the numbers of cases and deaths in late Autumn. Increasing the transmission rate by 20%, which is a hypothetical case of what can happen, should removing some further restrictions still currently in place result in a substantial growth in cases and deaths over the next few months, results in a very significant growth in cases and deaths that can even exceed those observed during the first epidemic wave. This highlights the importance of maintaining effective measures of social distancing and disease control when lifting quarantine restrictions.

**Figure 5.**
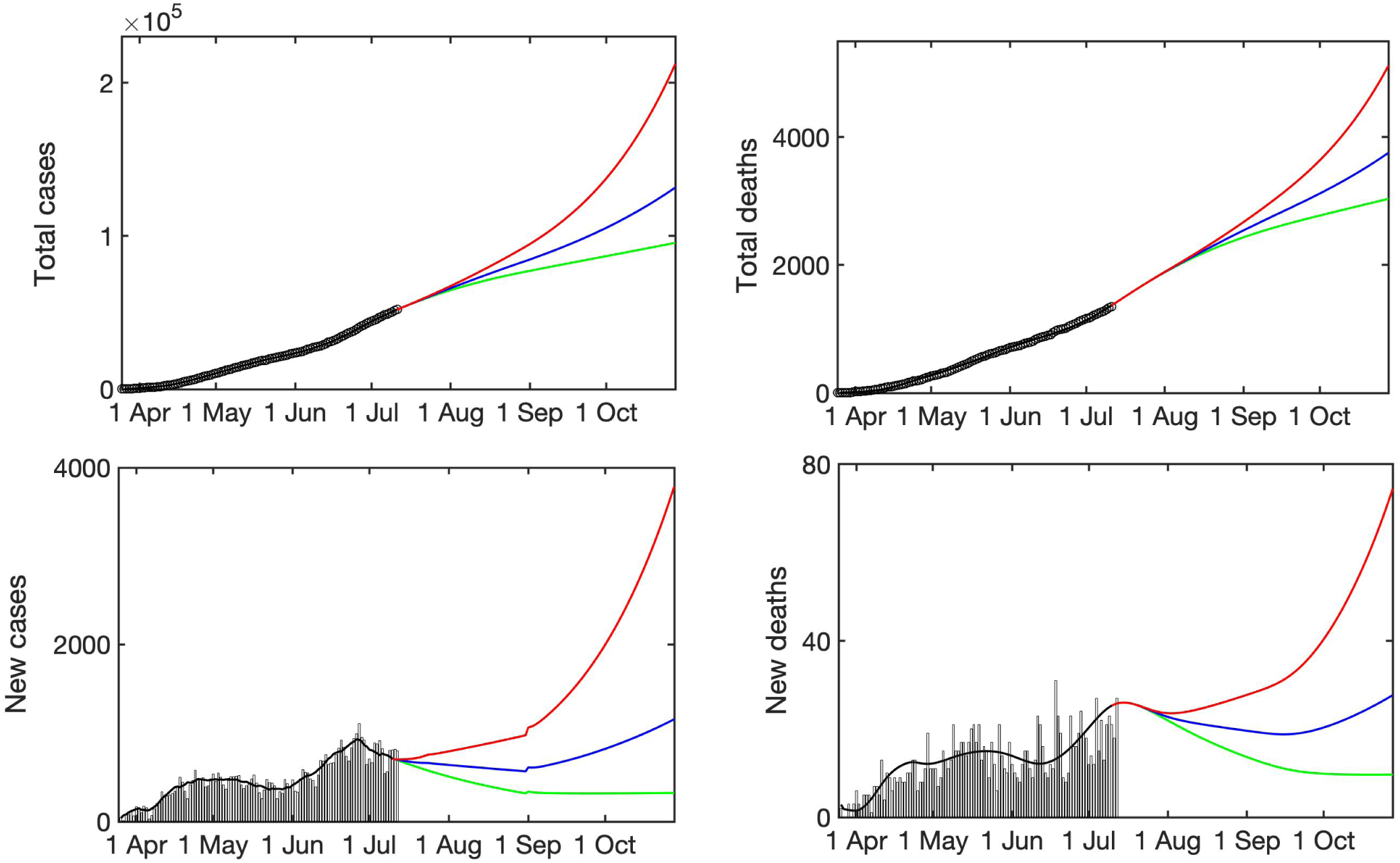
Longer-term forecast of epidemic dynamics of model (2) continuing with an overall transmission rate *β* as of 10 July 2020 (green), or the same rate being increased uniformly for all age groups by 10% (blue), and 20% (red) over initial two weeks, and then staying constant for the remaining duration of simulation.

To explore the potential impact of lockdown in the case where at baseline the infection is growing, in Fig. 6 we have fixed all parameter values as in Fig. 5, with a transmission rate increased by 20%, and compared a baseline, shown in red, when no lockdown is used, to three scenarios associated with a reduction in contacts between different age groups. The first scenario represented a 30% reduction in school contacts starting from the 1st September, which is the start of school and academic year in Ukraine. The second scenario models a 50% reduction in the level of contacts for people over 60, which represents an earlier policy of shielding that was in place from the 6 April until 5 June 2020. Compared to a restriction of mixing between school contacts, we observe a substantially smaller reduction in the number of total and daily cases, though a much less substantial change in the number of total and daily deaths, and on a longer timescale the reduction in school contacts proves to be more efficient in also reducing the daily and overall deaths. This can be attributed to a significant contribution of younger people to a spread of infection through their substantially larger numbers of contacts. The last scenario we consider is a 30% reduction in contacts among work contacts. We observe that this scenario represents the biggest reduction in both daily and total number of cases, but also, interestingly, over time it results in a smaller number of daily deaths, which eventually results in a smaller number of total deaths. This can be explained by a combination of mortality among 40-60-year olds, together with a reduction in the level of infection they are able to pass to older people, suggesting that promoting working from home and other ways of reducing work contacts appear to be the most efficient way of controlling the spread of infection in Ukraine, and minimising disease burden in terms of cases and deaths. This result is consistent with an earlier observation that working from home provided the largest contribution to reducing early cases of COVID-19 in Hong Kong^30^.

**Figure 6.**
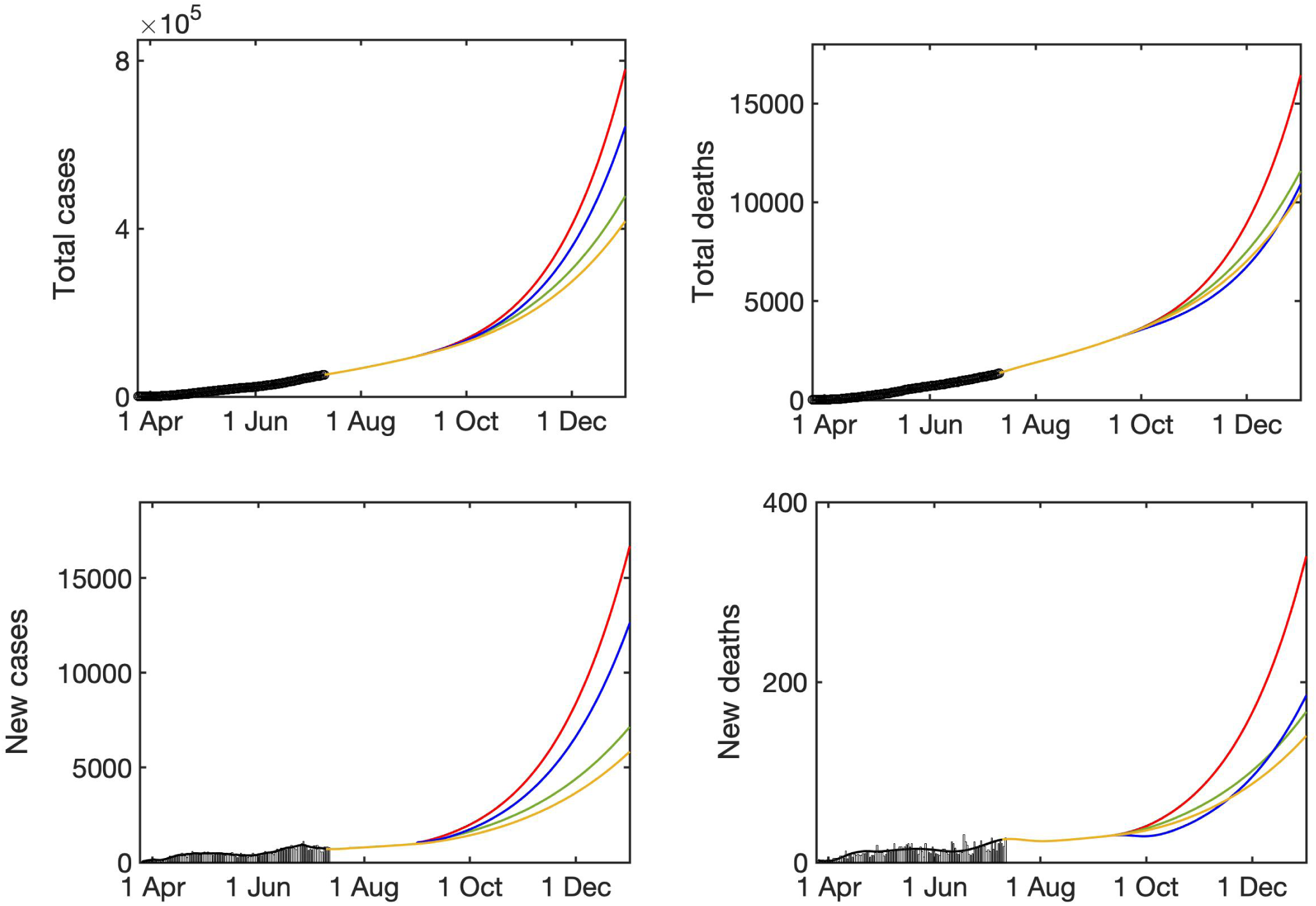
Modelling the effects of different quarantine strategies. Red line indicates the baseline forecast for a 20% increased transmission rate without any quarantine measures, green line denotes a 30% reduction in school contacts, blue line denotes a 50% shielding of over 60+ introduced gradually over two weeks, and light-brown line denotes a 30% reduction in work contacts. All intervention measures are introduced on the 1st September and are compared to a baseline shown in red.

## Discussion

In this paper we have developed and explored a mathematical model of COVID-19 in Ukraine, with an emphasis on including realistic distributions of characteristic times, population age structure and age-specific mixing patterns, as well as the impact of different lockdown scenarios. Our results indicate that in the short term, one should expect a continuing fast growth both in the number of infections, and the number of deaths. Current trend suggests that Ukraine has experienced the problem observed in many other countries: a lockdown was introduced very early into the outbreak, when the numbers of infected people were still relatively small, and then after 2-3 months, it was necessary to start lifting lockdown restrictions to reduce societal tensions and support the economy, but since by this point the prevalence was much higher, this quickly resulted in the high level of growth of new cases. Analysis of age distribution of cases and deaths reveals that after a short initial transient, by mid-April age distributions have reached stationary distributions, with subsequent growth in cases/deaths having almost no effect on these distributions. Whereas in most countries, the highest proportion of deaths is among over-70s, in Ukraine almost 50% of total deaths are in the age group 50-70, which can be partially attributed to a shorter life expectancy, but can also signify a rather high prevalence of co-morbidities that exacerbate the disease and result in death. The presence of co-morbidities and generally poorer health can also possibly explain another worrying observation concerning rather significant (compared to other countries) rates of death among younger age groups, such as 40-50, and even 30-40 years, which have almost no deaths in many other countries.

For the possible case of subsequent sharp growth in cases and deaths in Ukraine in the near future, we have analysed the effects different lockdown scenarios can have on these numbers. Quite naturally, we observed that restricting mixing among children and young adults has a higher impact on reducing the number of cases, due to higher mixing rates in these age groups, whereas shielding over-60s would have a smaller effect on reducing the number of cases, but would much more significantly reduce the number of deaths. Interestingly, the biggest reduction in both cases and deaths is associated with a moderate reduction in the level of contacts among the working-age population. This suggests that promoting working from home where possible, reducing travel to work by crowded public transport, and maintaining strict social distancing/prevention rules would, over longer time scale, reduce the number of cases, thus reducing the potential of hospitals being overwhelmed, and will also reduce the numbers of deaths.

There are several directions in which the work in this paper can be extended. One specific issue is having up-to-date and reasonably accurate mixing matrices for different types of contacts and different age groups in Ukraine. This could potentially by addressed using surveys that would elucidate patterns of contacts, as well as travel to work/shopping habits etc in a manner similar to the household survey being contacted since mid-March 2020 in the UK for an analogous work^31^. An alternative would be to try using results of simulations and short-term forecasting with the model to work backwards and deduce contact matrices that would provide the best fit for observed data. Another related issue concerns the fact that being Europe’s largest country by size, and sixth largest European country by population^32^, there is significant variation between different Ukrainian regions in terms of their population structure, local transportation modes, patterns of work etc, which naturally has a pronounced effect on how a disease is spreading in those regions. In this respect, conducting such surveys locally would allow one to much more accurately model the dynamics of local spread. Another interesting and important aspect that is not included in our model concerns the fact that besides an actual COVID-19 epidemic, we are currently experiencing an associated “infodemic”, whereby there is a large number of conspiracy theories about the virus being spread on social media^33,34^, which results in a notable proportion of population not believing in coronavirus and, as a result, not following social distancing guidelines, let alone using protective measures, such as face masks. This, of course, exacerbates the epidemic and makes it much harder to contain. Another direction, in which our model could be improved is to include the spread of disease awareness and compliance into a model^35,36^, which would also allow us to analyse different ways how to best target awareness campaigns. Finally, one the biggest challenges for any mathematical model of epidemics is the reliability of data, on which the model is based. Similarly to many other countries, in the specific context of COVID-19 in Ukraine, it is already known that there are some delays in collecting and reporting data, and improving this would result in more accurate parametrisation of the model, thus also improving its predictive power. In terms of SARS-Cov-2 testing, Ukraine is currently performing 14.9 tests per confirmed case, which is well within the WHO-recommended guideline of 10-30 tests per confirmed case as a benchmark of adequate testing^37^, though increasing testing capacity further may help identify and trace cases faster, thereby reducing the potential for disease spread.

## Methods

### Mathematical model

The SEIR-type mathematical model of COVID-19 in Ukraine has the form

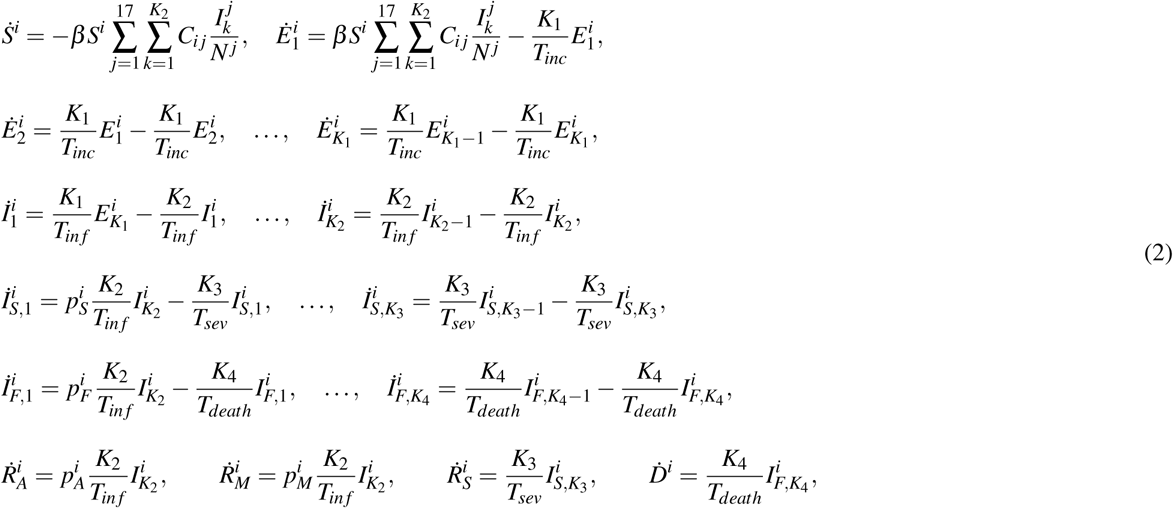

where *i* = 1..16 are different age groups, *N* ^*j*^ is the total population in *j*-th age group, *β* is the disease transmission rate, and *C*_*i j*_ is the mixing matrix given in (1). Age distribution of Ukrainian population according to the latest census data is illustrated in Fig. 7.

**Figure 7.**
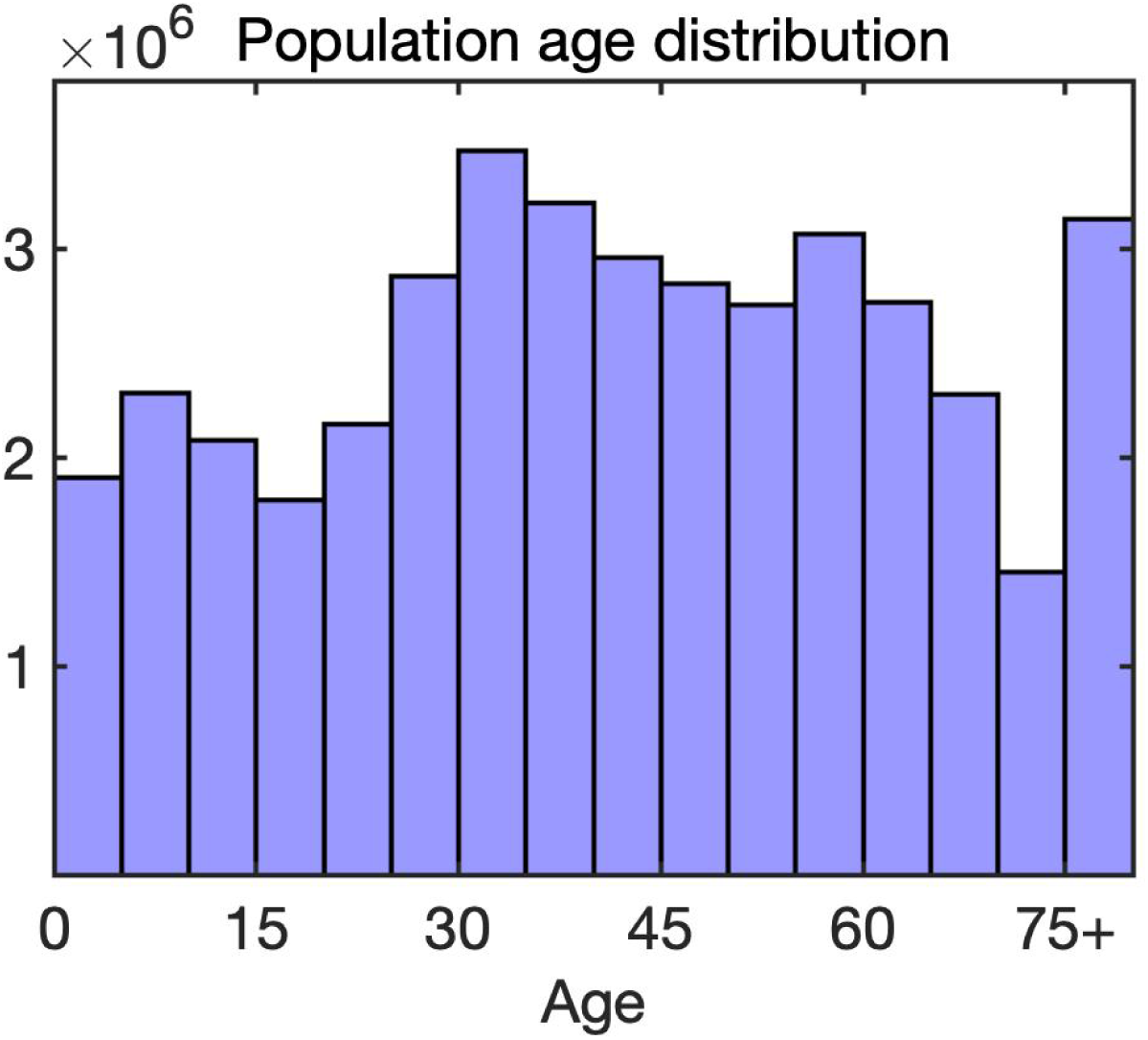
Population age distribution in Ukraine^49^.

### Mixing matrices

A POLYMOD project conducted in 2004-2008 collected data on age-specific contacts in various settings (all, physical, work, home) for eight European countries^38^, and more recently a BBC Contagion! program yielded more up-to-date data on contacts in the UK^39,40^. Using data from demographic and household surveys, Prem et al.^41^ have extended POLYMOD study and produced equivalent age-specific contact matrices for another 152 countries, and we have used the matrices for Ukraine from that study to represent contacts between individuals in our model. These matrices are illustrated in Fig. 8 for different location-specific settings.

**Figure 8.**
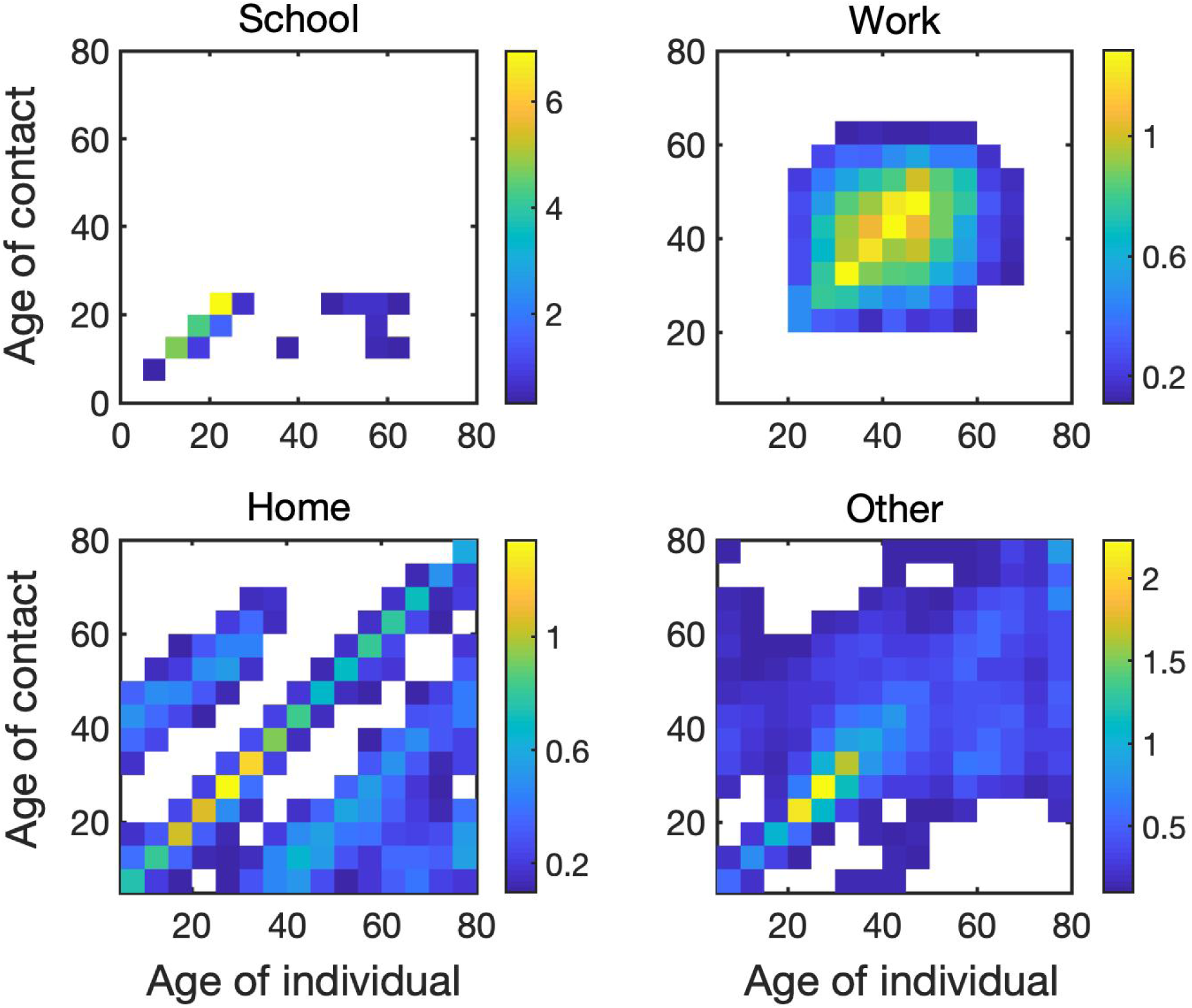
Age-specific and location-specific contact matrices for Ukraine^41^. The colour code denotes an average number of daily contacts between individuals in different age groups at each location.

### Parameter estimation

Age-stratified parameter values of proportions of asymptomatic and hospitalised individuals, as well as case fatality ratios, were provided directly by the Public Health Center of the Ministry of Health of Ukraine, and are contained in Table 1.

Basic reproduction number for the model (2) is given by the largest eigenvalue of *C*_*i j*_*N*^*i*^*/N* ^*j*^ multiplied by *β T*_*in f*_ ^42–44^. Despite the growth in the number of cases, since the total cases so far constitute around 0.1% of the total population of the country, the effective reproduction number can be well approximated by the basic reproduction number, as the population is still largely susceptible. We note that the latest data from European countries that have already undergone one full cycle of COVID-19 infection (i.e. a significant initial growth in cases, followed by the introduction of containment/lockdown measures, and a subsequent substantial reduction in cases and deaths), indicate that even at this stage the seroprevalence in those countries is still very low, in the region of 5-10%^45,46^. We have used a seven-day moving average of the number of reported cases (as established through RT-PCR testing) to obtain the number of new infections on any day, and then used this to obtain the value of *β* that was then used to simulate the dynamics for the next day, at which point the value of the disease transmission rate was again updated. This allowed us to follow very closely changes in disease transmission associated with the introduction and lifting of quarantine. The death rate was scaled to an aggregate value of 0.046 at day 30 after the start of a simulation by proportionally scaling all age-specific values of *p*_*F*_, and subsequently it was kept constant for the remainder of the simulation. The starting point for the simulation was chosen to be 25 March 2020, which is the date when the first 100 confirmed COVID-19 cases were reported.

## Data Availability

All data used in this study can be accessed directly through the dashboard of the Public Health Center of the Ministry of Health of Ukraine, with all parameter values provided in the text of the manuscript.

https://github.com/kblyuss/UKR_COVID

https://nszu.gov.ua/covid/dashboard

## Author contributions statement

All authors conceived the study, developed the model, performed numerical simulations, and analysed the results. All authors prepared and reviewed the manuscript.

## Additional information

### Data availability

All data used in this study can be accessed directly through the dashboard of the Public Health Center of the Ministry of Health of Ukraine^48^, with all parameter values provided in the text of the manuscript.

### Code availability

All codes used for simulations are available at https://github.com/kblyuss/UKR_COVID.

